# SIR Modeling the Dual Disaster Impacts of Omicron B.1.1.529 and Natural Disaster Events on Simulated 6 Months (December 2021 – May 2022) Healthcare System Resiliences in Fragile SE Asia Ring of Fire Ecosystems

**DOI:** 10.1101/2021.12.07.21267405

**Authors:** Andri Wibowo

## Abstract

For some countries that have experienced numerous natural disasters, including massive earthquakes and tsunamis, managing the COVID-19 pandemic can be very challenging. This situation arises considering that the disaster can directly and indirectly affect the healthcare system’ s capacity to serve the COVID-19 cases. With severely damaged healthcare facilities due to the disaster, there will be severely ill COVID-19 cases unmanaged. The coupling and interplay between these two phenomena can indeed be catastrophic. One of the regions where this issue becomes concerned is in Southeast Asia, where most of the Asian countries lie in the fragile ring of fire ecosystem, contributing to the high tsunami and earthquake disasters in the world. At the same time, Asia is one of the regions that have been severely impacted due to the current COVID-19 Delta Variant. Recently, a more contagious Omicron Variant has emerged and put a more massive burden on the healthcare facilities that are impacted by disasters. Then, in this situation, this paper aims to assess healthcare resilience in managing the Omicron pandemic amid disaster impacts. SIR simulation was used to determine whether severely ill Omicron cases were below or above healthcare and ICU capacity under different vaccination coverage. Our result confirms that vaccination coverage was the imminent factor in reducing the severely ill cases in every healthcare facility, whether the facilities were damaged or not. Increasing vaccination coverage from 30% to 60% will significantly reduce the number of severely ill cases that fall below the capacity of healthcare. Based on the current SIR model on the Omicron epidemic variables and Ro, it is estimated that the Omicron will reach its peak after 180 days in February 2022 and will totally disappear in May 2022 in this modeled area. When healthcare system facilities were fully operational and no disaster happened, combined with 60% vaccination rates, all Omicron case numbers were below and under the available hospital beds and even available ICU beds. While the situation is changed when a disaster occurs and causes 30% damage or reduction to healthcare facilities. In this situation, there are portions of Omicron cases that cannot be managed by the healthcare system since the cases have exceeded the available beds. The situations become more apparent where the healthcare facilities are severely damaged and lose 60% of their functionality. In this situation, all modeled Omicron cases and even the severe cases have exceeded the ICU capacity.

## 1. Introduction

### 1.1 Dual disaster potentials in SE Asia

The current COVID-19 pandemic is a complex global crisis without contemporary precedent. In just about every country around the world, the pandemic response is taking up the bulk of resources, expertise, time, and effort, causing an enormous burden on healthcare systems. However, how would people and healthcare systems cope if a major natural disaster, like an earthquake or a tropical cyclone, occurred simultaneously while the COVID-19 pandemic continued (Curkovic et al. 2021).

SE Asia is one of the most fragile ecosystems in the world. Its location, which lies on the ring of fire, has caused most of the countries in Southeast Asia to have frequent natural disasters recently, ranging from tsunamis to earthquakes. The occurrence of those massive disasters has caused massive casualties due to natural disaster impacts but has also caused casualties due to communicable diseases that emerged after the disasters (Şimşek & Gunduz 2021). The following are examples of epidemics following natural disasters.

The South Asian Tsunami in 2004that killed more than 250,000 people and displaced more than 1.7 million across 16 countries has produced ideal conditions for an acute respiratory infection outbreak in Aceh, Indonesia, the worst hit region. In 2010, the first cholera outbreak in more than a century in Haiti resulted in 8,183 deaths, which was amplified by damage to infrastructure caused by the preceding earthquake.

In fact, following any meteorological event, ranging from cyclones, floods, to tornadoes or geophysical events, including earthquakes and volcanic eruptions, that has displaced large numbers of people, has led to the emergence of epidemic diseases, including diarrhoeal diseases, hepatitis A and E, leptospirosis, measles, meningitis, acute respiratory infections, malaria, and dengue.

### 1.2 Disaster impacts on healthcare systems

Dual disasters have happened and are related to the collapse of healthcare systems and systems. The collapse of the healthcare system was related to these direct and indirect causes. The direst causes are the healthcare system infrastructures in the form of hospital buildings and/or related healthcare support infrastructures that are damaged due to the tsunami and earthquake. While the indirect causes are the support systems of healthcare systems that were affected, this included the damage to road networks. This is considering that road networks are used to support the logistical distribution of healthcare equipment, medicines, and also the remobilization of healthcare staff.

This situation has been reported recently, taking the tsunami disaster as an example. As reported by Suwandono et al. (2005), the previous tsunami in 2004 reduced the capacity of healthcare systems from 50% to almost 80%. In detail, 43% of community health facilities are heavily damaged and only 25% are less damaged. Besides that, healthcare human resources have been reduced by up to 40%. As a comparison, similar situations were also observed in the post-earthquake in Bantul. As reported by Widayatun and Fatoni (2013), approximately 55.6% of healthcare facilities were damaged.

### 1.3 Omicron

On November 26, 2021 (WHO 2021), WHO designated the SARS-CoV-2 Variant B.1.1.529 as a Variant of Concern (VoC) based on advice from WHO’ s Technical Advisory Group on Virus Evolution (VOC). This variant has been given the name Omicron. South Africa reported the Omicron to WHO for the first time on November 24, 2021. In South Africa, infections were high in recent weeks, coinciding with the discovery of Omicron. The first known confirmed Omicron infection came from a specimen collected on November 9, 2021, and the first publicly available sequence came from a specimen collected on November 11, 2021. The number of cases of this variant appear to be on the rise in a number of South African provinces. The Omicron was also discovered in Botswana in samples collected on November 11, 2021. As of November 28, 2021, at 3 PM, cases had been detected in four additional countries.

Omicron has some deletions (Karim and Karim 2021) and more than 30 mutations, some of which overlap with those in the alpha, beta, gamma, or delta VoCs, including 69–70del, T95I, G142D/143–145del, K417N, T478K, N501Y, N655Y, N679K, and P681H. These deletions and mutations are known to increase viral transmissibility, viral binding affinity, and antibody escape. Some of the other known omicron mutations confer increased transmissibility and affect binding affinity (Greaney et al 2021, Harvey et al. 2021).

### 1.4 Case studies

Among five islands, Java is one of the most populated islands that have the most fragile ecosystems. This fragility is related to the geological characteristics of Java Island. Related to this, the Java Island landscape consists of numerous volcanic mountains that are still active. Even on December 4, 2021, Mount Semeru in East has erupted. Aside from volcanic mountains, Java Island is also vulnerable to tsunamis. Imminent causes of tsunamis on Java’ s island coasts are related to the presence of Krakatau, an active volcano in West Java. As a result, in December 2018, Krakatau erupted and caused a tsunami on the nearby coastline. The most heavily impacted coast due to the tsunami caused by the Krakatau eruption is the South West Coast of West Java.

Besides being prone to natural disasters, the Java Island has the highest COVID-19 confirmed cases. The record was observed in the period of June–August 2021 due to the COVID-19 Delta Variant. At that time, the surge of COVID-19 cases caused a nationwide bed occupancy rate (BOR) to reach 69%, which exceeded the WHO level of 60%. Java Island recorded the most BOR with a range of 71%-90%. This BOR was higher than islands outside the Java Islands, which ranged from 61% to 64%.

## 2. Methods

### 2.1. Modeled Area

The modeled area was a fragile ecosystems located in the South West Coast of West Java between 6°1’ 0’’ – 6°9’0’’ S and 105°2’0’’ – 106°0’0’’ E. This area was experiencing massive tsunami in the last 2018. The magnitude and damage caused by tsunami can be seen in Table 1.

**Table 1.**
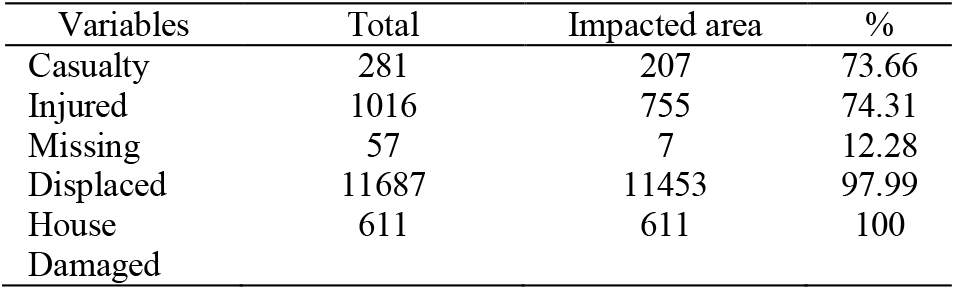
2018 tsunami’ s casualties and damages in modeled and impacted area

### 2.2. Demography of modeled Area

The modeled area was governed by a district level government. Then this area was populated and supported by healthcare system including hospitals. The demography data along with healthcare facilities including, vaccination rate, numbers of available beds, and Integrated Care Units (ICU) were available in Table 2.

**Table 2.** Demography and healthcare facility in modeled area

| Variables | Total |
| --- | --- |
| Populations | 1,214,000 |
| Vaccination | 22.02% |
| Hospitals | 4 |
| Beds | 302 |
| ICU | 21 |

### 2.3 SIR modeling

A well-known Susceptible Infectious Recovered (SIR) model was used in this study (Jo et al. 2020, Zhao et al. 2020). The SIR is regarded as a versatile compartmental mathematical tool for modeling any pandemic dynamic in dealing with the current COVID-19 pandemic. The model aids us in comprehending the kinetics of a pandemic in any specific aspect, and the SIR model is also simple to understand and has clear interpretations (Wagh et al. 2020).

The total population (N) is divided into three compartments in the standard SIR model (Waqas et al. 2020). The Susceptible (S), a subset of the total population that is vulnerable and at risk of infection, The Infected (I) are people who have been infected. The recovered (R), which is the percentage of the total population that recovers, Several input parameters must be considered in the SIR model. These parameters are denoted as ß.

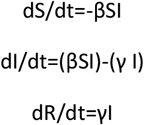

### 2.4 SIR variables

SIR consists of epidemic, healthcare, and demography variables (Table 2). The epidemic variable consisted of the reproductive numbers (Ro) of Omicron and vaccination rates. While the healthcare and demography variables consist of the number of available beds and ICUs, The model ran from December 2021 to May 2022. The period was based on people mobilizing for the end-of-year holiday and warnings from meteorological and climatological agencies about a possible tsunami due to extreme weather at the end of December 2021.The 6 month period was chosen considering that the reconstruction of healthcare facilities requires 6 months.

The SIR was implemented at several vaccination rates and in the capacity of healthcare facilities. The vaccination rates ranged from 30%, 60%, and 90%. Similar rates were applied to healthcare capacities. The available functional healthcare capacities were following the disaster impacts with the lowest capacity (30%) represents the severe impacts and damages.

## 3 Results and Discussion

The results of simulated Omicron-related infectious and severely ill cases related to vaccination coverage and healthcare conditions are available in Figure 1-3. The infections and severely ill cases were simulated according to the conditions of healthcare facilities due to the damage caused by natural disasters like tsunamis and earthquakes. From the results, first, it is obvious that vaccination is the most important factor in reducing the Omicron cases.

**Fig 1.**
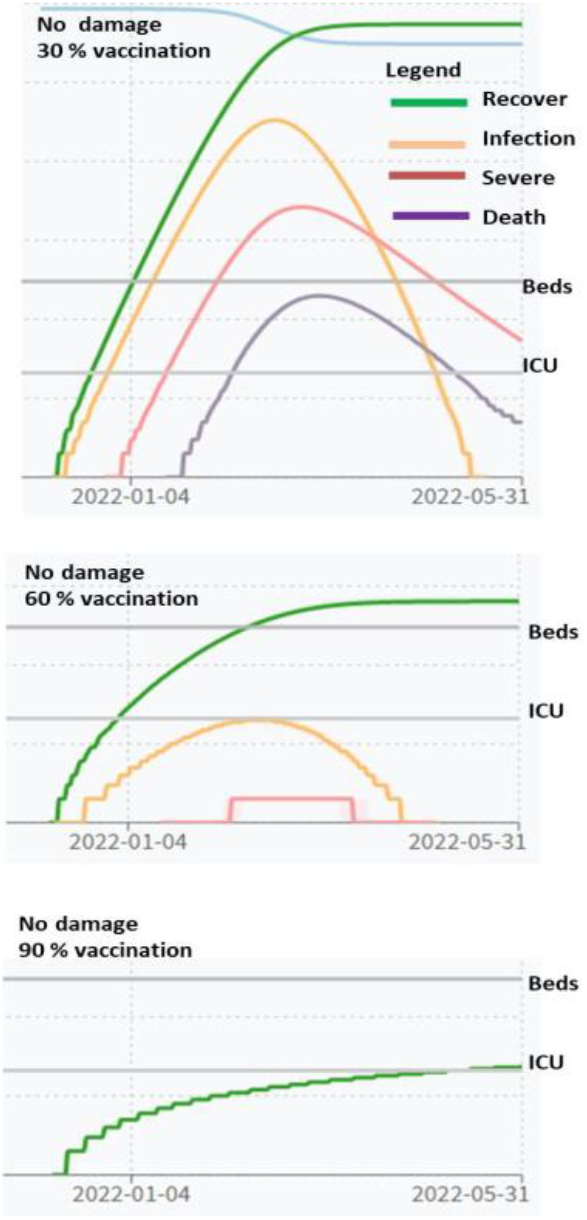
Omicron SIR model with no healthcare damages and vaccination rates of 30%, 60%, and 90%.

**Fig 2.**
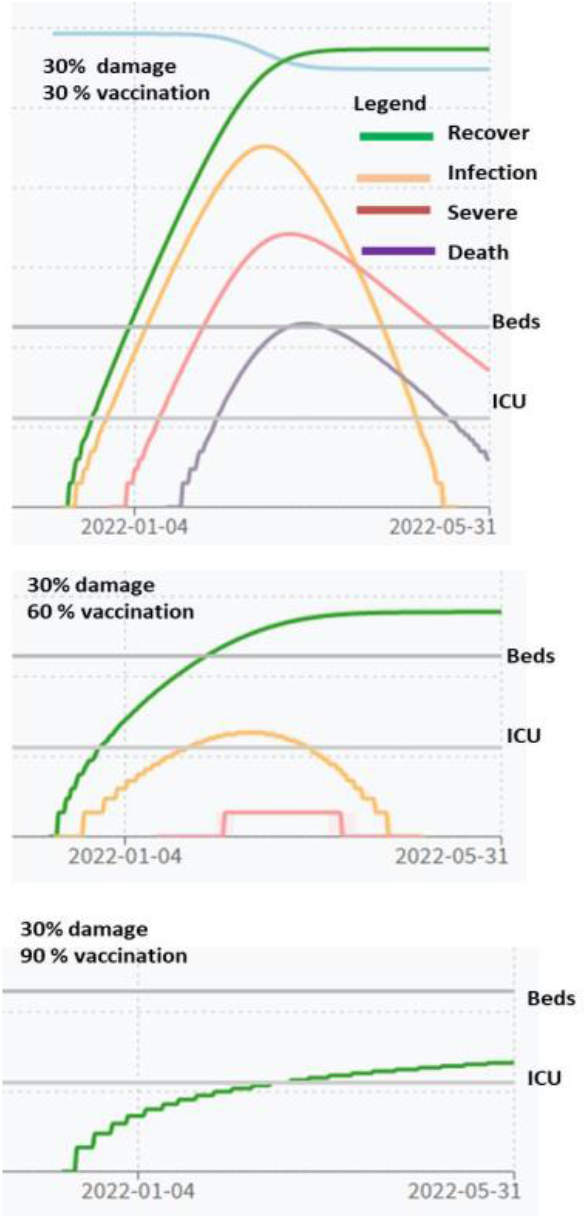
Omicron SIR model with 30% healthcare damages and vaccination rates of 30%, 60%, and 90%.

**Fig 3.**
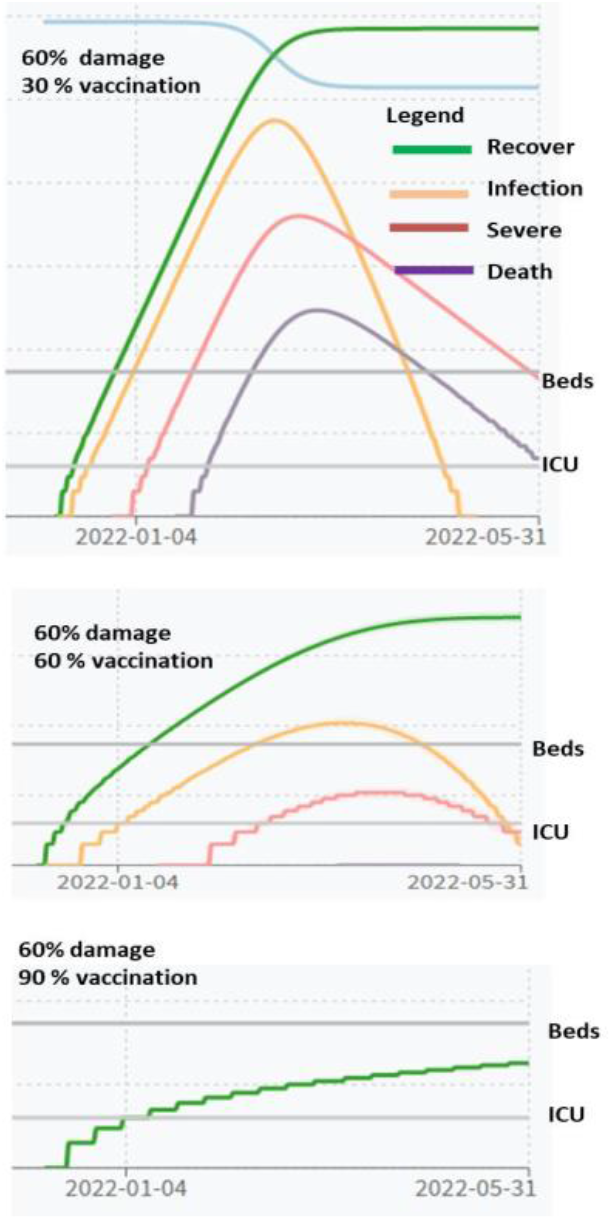
Omicron SIR model with 60% healthcare damages and vaccination rates of 30%, 60%, and 90%.

These facts were related to the conditions where, in every healthcare facility with damage rates of 0–60%, the Omicron cases were always reduced to zero with vaccination coverage close to 90%. While in almost every healthcare situation, the severely ill cases were always above healthcare facility capacities if only 30% of the population got the vaccine. By increasing the vaccination coverage up to 60%, all severely ill cases were below healthcare facility capacities and under control.

Currently, the presence of multiple mutations of the spike protein in the receptor-binding domain suggests that Omicron may have a high likelihood of immune escape from antibody-mediated protection. However, immune escape potential from cell-mediated immunity is more difficult to predict. Overall, there are considerable uncertainties in the magnitude of immune escape potential of Omicron.

Further research is needed to better understand the escape potential against vaccine- and infection induced immunity. Research efforts are ongoing, and the data are expected to be available in the coming weeks. Despite uncertainties, it is reasonable to assume that currently available vaccines offer some protection against severe disease and death.

The SIR model is considered a very useful tool and approach to estimate where a certain epidemic will reach its peak. Based on the current SIR model on the Omicron epidemic variables, it is estimated that the Omicron will reach its peak after 180 days in February 2022 and will totally disappear by May 2022. This result is in agreement with other COVID-19 variant SIR models. In comparison, the Delta variant, which is related to Omicron variants, reached its peak after 18 days. Using Ro close to 1.8, Saldana and Velasco-Hernandez (2021) found that the simulated SIR model of Delta reaches its peak on the 200th day. While the time when the Omicron will reach its peak depends on the Ro values, As Saldana and Velasco-Hernandez (2021) confirm, with Ro increasing from 1.8 to 5.0, there is a great possibility that the Omicron can reach its peak just within 20 days.

The most important finding in this study is how the disaster can affect the health facility’ s performance in mitigating the Omicron cases. The impacts were clearly visible where different damage levels were applied. When healthcare system facilities were fully operational and no disaster happened, combined with 60% vaccination rates, all Omicron case numbers were below and under the available hospital beds and even available ICU beds. It means that the Omicron cases can be managed since they are not surpassing the healthcare capacities (Fig.1). While the situation is changed when a disaster occurs and causes 30% damage or reduction to healthcare facilities. In this situation, there are portions of Omicron cases that cannot be managed by the healthcare system since the cases have exceeded the available beds. The situations become more apparent where the healthcare facilities are severely damaged and lose 60% of their functionality. In this situation, all Omicron cases and even the severe cases have exceeded the ICU capacity (Fig. 2).

Our results indicating the healthcare facilities lose its capacity to deliver healthcare service is in agreement with previous study. Following Camp Fire events, the continued closure of the Feather River Hospital after the Camp Fire resulted in a 17% reduction in the total number of staffed beds in Butte County. While the patient demand for hospitals increased due to the loss of 100 staffed beds because of damage on healthcare facilities. The challenges are exacerbated when pandemics are combined with natural disasters, especially if the natural disaster has a relatively short return period, such as tsunami and earthquake. The occurrence of both events concurrently or spatially over time can have disastrous consequences for community infrastructure, social, and economic institutions, necessitating prompt and timely prevention and control measures.

Healthcare systems, in particular, play a critical role in minimizing losses and saving as many lives as possible. When both events occur, managing healthcare systems may be difficult due to the various conflicting mitigation strategies required for each event separately. (Hassan & Mahmoud 2021, Kouadio et al 2012).

## Conclusions

Dual disasters are inevitable, and they require versatile tools to estimate the potential impacts of disasters on pandemic mitigation. The SIR model is one versatile tool that can assess how the loss of healthcare systems and functionality due to disasters can significantly reduce the number of COVID-19 cases that can be managed. This situation becomes more severe when dealing with the COVID-19 Varian that has high transmissibility, including the latest Omicron B.1.1.529 Variant.

## Data Availability

All data produced in the present work are contained in the manuscript

